# Developing an OMOP-Standardized Prostate Cancer Database and Improving Data Quality Using NLP and PSA-Based Algorithms

**DOI:** 10.64898/2026.06.30.26356984

**Authors:** Jiefei Wang, Jamaal C. Jackson, Anette Garza, Suhas Nalla, Tammy Adams Ninnemann, Yuanyi Zhang, Yong-Fang Kuo

**Author notes:** Corresponding Author: Jiefei Wang, PhD; 1005 Harborside Drive, Galveston, TX 77555, USA.

## Abstract

**Objective:** To develop and evaluate an Observational Medical Outcomes Partnership-(OMOP-) standardized prostate cancer database from the University of Texas Medical Branch (UTMB) Epic Electronic Health Record (EHR) and improve data quality using natural language processing(NLP) and prostate-specific antigen-(PSA) based algorithms.

**Materials and Methods:** We built a data pipeline to transform UTMB Epic EHR data for patients identified through the Galveston Cancer Registry (2010-2021) into OMOP Common Data Model (CDM) v5.4. Data quality was assessed by comparing the OMOP-standardized data with Galveston Cancer Registry data using availability agreement, Cohen’s kappa and Intraclass Correlation Coefficient. NLP was used to extract PSA, Gleason score, and cancer stage from clinical text, and PSA-based algorithms were used to identify missing treatment and biochemical recurrence.

**Results:** We extracted 815 analytic cases from UTMB EHR. Among them, 700 (85.9%) were complete and concordant with the cancer registry. PSA showed excellent value agreement. Structured Gleason score and stage data were sparse (n<20), but NLP greately improved capture. Treatment agreement was good compared with the cancer registry and improved slightly for radical prostatectomy after applying a PSA-based algorithm. Using PSA trajectories, we identified 60 cases of biochemical recurrence.

**Discussion:** The OMOP-standardized data from UTMB showed good agreement with the cancer registry. However, structured EHR fields incompletely captured diagnosis, pathology, and treatment details. NLP and PSA-based algorithms substantially improved data capture. Manual review also revealed errors in registry data, showing that OMOP-standardized EHR data can complement and help improve cancer registry quality. **Conclusion:** OMOP standardization combined with NLP and PSA-based algorithms improved prostate cancer data quality and research readiness.

**Lay Summary:** Electronic health record (EHR) systems contain valuable long-term health information about cancer, but their use is limited by difficulties in data extraction, the lack of standard data formats, and missing information in structured fields. In this study, we built a data pipeline to convert EHR data into the Observational Medical Outcomes Partnership (OMOP) Common Data Model, a widely used standard format that facilitates data sharing and collaborative research. To address missing data, we used natural language processing (NLP, a computational technique that enables computers to interpret human language) to extract prostate-specific antigen (PSA) levels, Gleason scores (a measure of cancer aggressiveness), and cancer stages from unstructured clinical notes. We also applied algorithms that track PSA blood trends over time to identify unrecorded cancer treatments and detect cancer recurrence. Comparing this database with a cancer registry demonstrated that standardized EHR records provide accurate clinical information and long-term follow-up. While structured EHR fields lacked key tumor characteristics, NLP dramatically improved their capture. Tracking PSA trends identified previously unrecorded treatments and flagged 60 potential cancer recurrences. Overall, combining standardized EHR formats with NLP and PSA-based algorithms creates a rich database to support real-world prostate cancer research.

## INTRODUCTION

Prostate cancer is the most commonly diagnosed non-skin cancer among men in the United States. The American Cancer Society projects that approximately 333,830 new cases of prostate cancer will be diagnosed in 2026, and more than 3.5 million men in the United States are currently living with prostate cancer^2^. Despite its high prevalence, prostate cancer generally has a favorable prognosis, with a 5-year relative survival rate approaching 98%. Consequently, long-term surveillance is essential for monitoring disease progression, treatment patterns, and outcomes among prostate cancer patients.

Electronic health record (EHR) data have increasingly been used to generate real-world evidence in prostate cancer^4,9,16^. Several studies have leveraged institutional EHR systems to examine prostate-specific antigen (PSA) trends, treatment effectiveness, and survivorship outcomes among prostate cancer patients^7,16,19^. Compared with traditional cancer registries, EHR data provide richer clinical detail and enable the capture of longitudinal patient information over time^1,5^. These characteristics make EHR systems a valuable resource for studying disease trajectories and monitoring treatment responses in prostate cancer populations.

Despite these advantages, several challenges limit the use of institutional EHR data for rigorous research^9^. First, EHR systems are primarily designed for clinical documentation and billing rather than research purposes. As a result, their structured tables may not capture key clinical variables, and important information (e.g., Gleason score details, pathology findings, and clinical assessments) can be embedded within unstructured clinical notes. Second, the lack of standardized data structures across institutions complicates data sharing and interoperability. Consequently, many studies relying on institutional EHR data focus primarily on structured baseline variables, thereby overlooking the broader capabilities of EHR data for capturing longitudinal outcomes and valuable clinical variables.

To maximize the research potential of EHR data, there is a critical need for a standardized data framework. The Observational Medical Outcomes Partnership (OMOP) Common Data Model (CDM) is one of the most widely adopted framework for organizing healthcare data using standardized structures and vocabularies^8^. Standardizing institutional EHR data within the OMOP framework facilitates reproducible analyses and supports interoperability across institutions. Although OMOP CDM has been widely adopted for observational health research and increasingly used in oncology, relatively few studies have focused on developing prostate-cancer-specific OMOP datasets^17^, and even fewer have integrated structured EHR data with information extracted from unstructured clinical notes^20^.

This study aims to develop an OMOP-standardized prostate cancer database at the University of Texas Medical Branch (UTMB), referred to as the UTMB-OMOP database, and establish a pipeline to transform UTMB Epic EHR data into the OMOP CDM. We evaluated UTMB Epic EHR data across key points in the prostate cancer care continuum. To address data limitations, we used PSA-based algorithms to detect missing treatment and biochemical recurrence (BCR); We also applied natural language processing (NLP) to extract PSA, Gleason score, and cancer stage information from unstructured clinical notes. This study contributes by: (1) developing a pipeline for constructing an OMOP-standardized prostate cancer database from Epic EHR; (2) implementing several PSA-based algorithms for improving database completeness and usability; and (3) evaluating whether NLP improves the completeness of PSA, Gleason score, and cancer stage data, and assessing agreement between the extracted values and cancer registry data. The resulting data pipeline and analytic framework will provide a reproducible model for other institutions seeking to standardize Epic EHR data and support large-scale prostate cancer research.

## 1 METHODS

### 1.1 Patient Cohort

The data for this study were sourced from the UTMB EHR. The study was approved by the UTMB Institutional Review Board (Record Number: 23-0223). Prostate cancer patients were identified using data from the UTMB-Galveston Cancer Registry (GCR) for diagnoses between January 1, 2010, and December 31, 2021. The GCR patients included two distinct case types: analytics cases and non-analytics cases. Analytic cases were those patients who were diagnosed and/or received their first course of treatment at UTMB. Non-analytics cases were those patients who were diagnosed and received their first course of treatment at another institution but had subsequent care at UTMB. We restricted our cohort to analytics cases only since UTMB serves as the primary reporting source for analytics cases. The patients’ medical record numbers (MRN) were used to extract their corresponding EHR data from UTMB’s Epic system. Patients were excluded who were (1) younger than 18 years at the time of diagnosis or (2) incarcerated within the Texas Department of Criminal Justice (TDCJ) and received care at UTMB.

### 1.2 OMOP Data Pipeline

We developed an Extract, Transform, and Load pipeline to convert UTMB Epic EHR data into the OMOP CDM v5.4 format. Our pipeline specifically focused on 12 clinical tables crucial for prostate cancer research (See Supplemental Table 1 for their definitions). The pipeline involved three main stages, all implemented using R^14^. First, vocabulary mapping utilized a hybrid approach: standard clinical coding systems (like ICD-10-CM, CPT4, LOINC) were mapped using Athena (official repository of OMOP standard vocabularies), while local codes in the person table were manually mapped with USAGI^12^ by an experienced biostatistician. Second, table-level transformation converted source EHR tables into their corresponding OMOP CDM formats. Finally, the transformed data were aggregated into the appropriate OMOP CDM tables. To ensure data completeness, we retained records from all relevant source tables, even when duplicates existed (e.g. the same diagnosis code can appear multiple time due to documentation and billing needs). The details of the source tables, transformation rules, and destination OMOP CDM tables are described in Supplemental section 2.

To evaluate the OMOP CDM transformation quality, we reported the number of records and concepts mapped to standard OMOP concepts within each OMOP CDM domain. Percentage mapping rates were calculated as the number of records mapped to standard concepts divided by the total number of records in the source EHR dataset. For a table with low mapping rate, we reviewed the most common unmapped concepts to identify reasons for mapping failure and to assess whether they were relevant to prostate cancer research.

### 1.3 Data Quality Assessment

Figure 1 illustrates the cancer care continuum. It represents the patient trajectory from diagnosis and evaluation to treatment, surveillance, and potential recurrence. These stages are important for cancer research. We evaluated our OMOP CDM data quality at each critical stage. To compare data completeness, we used availability agreement. To compare value agreement, we used Cohen’s kappa (κ) for categorical variables, and Intraclass Correlation Coefficient (ICC) for numerical variables. For availability agreement, we defined values < 0.70 as poor, 0.70−0.85 as moderate, 0.85−0.95 as good, and > 0.95 as excellent agreement. For κ statistics, values ≤0.40 were considered poor, 0.41−0.60 moderate, 0.61−0.80 good, and ≥ 0.80 excellent agreement. For ICC, values < 0.50 were considered poor, 0.50−0.75 moderate, 0.75−0.90 good, and > 0.90 excellent agreement^10,11^.

**Figure 1.**
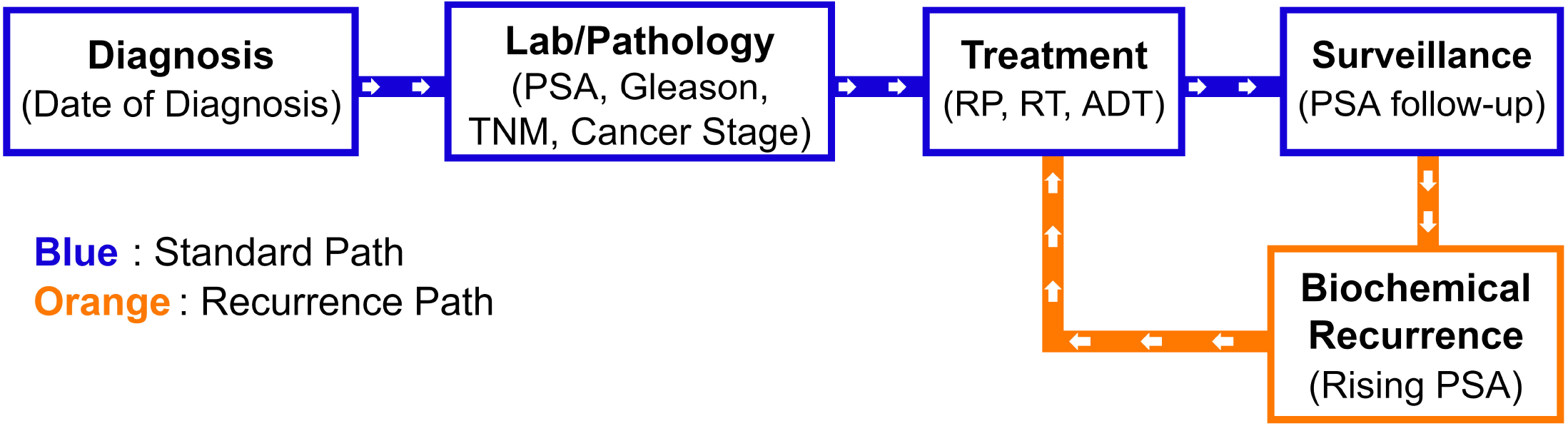
Cancer Care Continuum. PSA: prostate-specific antigen; RT: radiation therapy.

#### 1.3.1 Diagnosis Assessment

For the prostate cancer dataset, it is important to ascertain the date of initial diagnosis. Therefore, we compared the diagnosis dates of prostate cancer patients between the UTMB-OMOP and the GCR cohort. In the UTMB-OMOP, we used the standard concept “Neoplasm of prostate” from the condition_occurrence table to identify the date of diagnosis. Patients were considered incomplete if they had no recorded prostate cancer diagnosis in the UTMB-OMOP. We calculated the time difference between the UTMB-OMOP and GCR cohorts. Because diagnosis dates can differ between systems due to variations in documentation practices (e.g., waiting for a pathology report before giving an ICD code) and delays in coding and registry abstraction, we allowed a 4-month window for concordance. Patients were flagged as discordant if diagnosis date differences were larger than four months. We reported the number of incomplete and discordant patients, then reviewed these cases to identify reasons for discrepancies. In this study, clinical chart review was conducted by Suhas Nalla, a medical student, under the supervision of Jamaal C. Jackson, MD, an attending urologist. Any ambiguous cases were referred to Dr. Jackson for clinical adjudication and data verification. For all subsequent analyses, we restricted our analysis cohort to complete and concordant patients. The true diagnosis date was calculated as the earlier date between the UTMB-OMOP and GCR.

#### 1.3.2 Lab and Pathology Assessment

We compared the completeness of PSA level, Gleason score, and cancer stage group (AJCC 8th Edition^13^) between the UTMB-OMOP and the GCR cohort. ICC was used to assess agreement for log-transformed PSA levels and Gleason scores, while Cohen’s κ was used to evaluate agreement for cancer stage group. For all comparisons, we focused on clinical stage and constrained the reporting time window to range from 30 days before the diagnosis date through the earlier of the initial treatment date recorded in GCR or 365 days after diagnosis (365 days after diagnosis if no treatment was documented). This time window was selected to match the GCR reporting criteria^18^. Since a patient can have multiple PSA values, we used the maximum PSA for comparison. For value agreement in cancer stage, we dichotomized stages into early (Stages I-II) and late (Stages III-IV). Since GCR began recording PSA, Gleason score, and AJCC cancer stage in 2018, we restricted the analysis to patients diagnosed in 2018 or later.

Additionally, we extracted PSA, Gleason score, and AJCC TNM stage data from clinical notes using the Gemma 4 large language model (LLM). Stage group was calculated from PSA, Gleason grade group, and TNM stage (see details in the supplemental section 3). The extracted variables were stored independently of the UTMB-OMOP database and combined during analysis phase. We compared the structured data before and after augmentation with NLP-extracted values against the corresponding GCR records, using the same agreement metrics and inclusion criteria described above for the UTMB-OMOP cohort.

#### 1.3.3 Cancer Treatment Assessment

To evaluate treatment patterns and outcomes among patients with prostate cancer, we identified individuals in the UTMBOMOP who received radical prostatectomy (RP), radiation therapy (RT), or androgen deprivation therapy (ADT). Because treatment records in UTMB-OMOP do not explicitly indicate line of therapy and the GCR dataset captures only the first course of treatment, we aligned treatment data in UTMB-OMOP to GCR by restricting records to those occurring between 1 month before and 1 year after the diagnosis date. This time window was selected based on clinical rationale to approximate first-course treatment, as definitive therapy for prostate cancer is typically initiated within the first year following diagnosis. However, it should be noted that this restriction mitigates but does not eliminate potential misclassification, as some treatments occurring within 1 year may represent salvage or subsequent therapy rather than true first-course treatment. All analyses were conducted for descriptive purposes.

Because some patients received treatment outside our institution, leading to incomplete capture in UTMB-OMOP, we applied previously developed PSA-based missing-treatment detection algorithms^3^ to identify potentially missing treatment records. We implemented the algorithms in R based on the published pseudocode. Complete rules, data handling, and adaptations are provided in Supplementary Methods Section 6. Treatment patterns were compared before and after applying the missing-treatment detection algorithms. Finally, we manually reviewed all patients flagged by the algorithms to identify sources of misclassification.

#### 1.3.4 Recurrence Assessment

Prostate cancer recurrence is a critical outcome measure for assessing treatment efficacy and long-term patient prognosis. We captured BCR events based on PSA values. For patients who underwent radical prostatectomy, we used the American Urological Association definition^6^. BCR is defined as a PSA value ≥ 0.2 ng/mL followed by a subsequent PSA that also remains ≥ 0.2 ng/mL. For patients who received radiation therapy, BCR is defined using the Phoenix criterion^15^. A PSA rise of more than 2 ng/mL above the post-treatment nadir is considered as BCR. The nadir is determined as the absolute minimum PSA observed after treatment. If multiple BCRs were found, the first BCR was used for analysis. We reported the number of BCR and median time (in months) to the BCR event. Additionally, we manually reviewed those patients with BCR to calculate the positive predictive value (PPV) of the algorithm. For the manual review, we flagged patients as having true recurrence when any of positron emission tomography (PET), magnetic resonance imaging (MRI), computed tomography (CT), or bone scans shows positive results.

## 2 RESULTS

Between 2010 and 2021, UTMB reported to GCR 1865 prostate cancer records for patients aged 18 or older. Figure 2 illustrates the data flowchart. After excluding two duplicate MRN records, 688 incarcerated patients, six patients without EHR records, and five patients with identity mismatches between registry and EHR records, we identified 1164 patients with prostate cancer from UTMB EHR. Table 1 summarizes the number of records and concepts mapped to standard OMOP concepts in each OMOP CDM domain. Among them, the condition_occurrence and measurement tables had the highest mapping rate (99.46% and 92.76%, respectively), while the procedure_occurrence and visit_occurrence tables had the lowest mapping rates (73.55% and 67.86%, respectively). The reason for the unmapped concepts was primarily the presence of local codes that had no corresponding standard OMOP concepts. For visit_occurrence, the most common unmapped local codes were “historical encounter” (9.86%) and “orders only” (7.47%). For procedure_occurrence, the most common unmapped local codes were “faculty present with patient” (3.16%) and “EKG 12-Lead” (2.35%). Those codes are important to clinical workflow, but not directly relevant to cancer research. These results highlight the difference between healthcare and research data needs.

**Figure 2.**
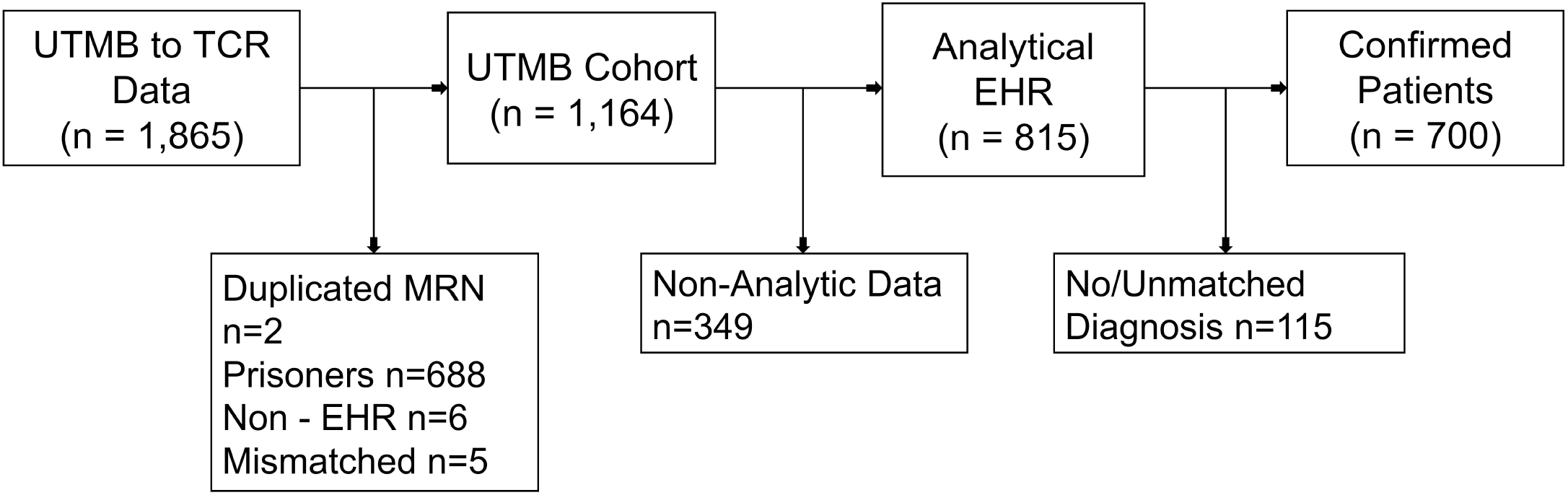
Cancer Database. UTMB: University of Texas Medical Branch; TCR: Texas Cancer Registry; EHR: Electronic Health Record; MRN: medical record number.

**Table 1.**
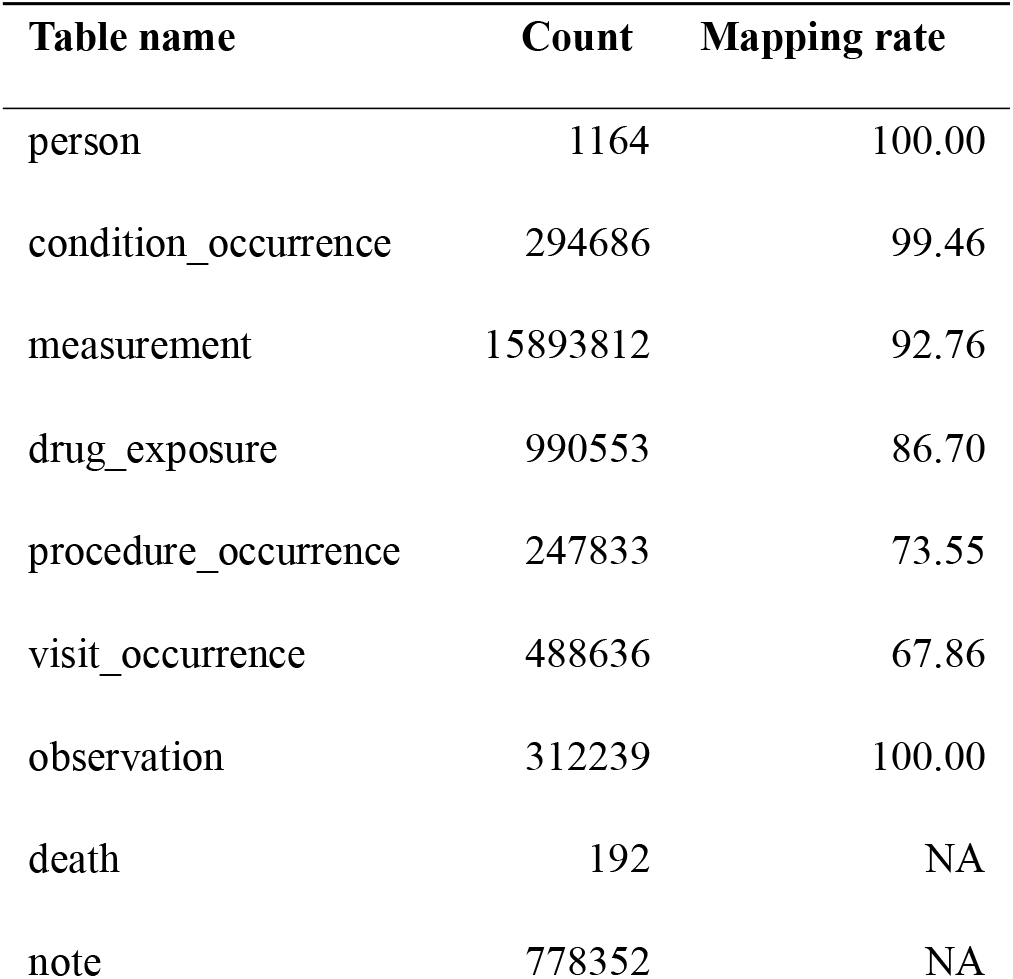
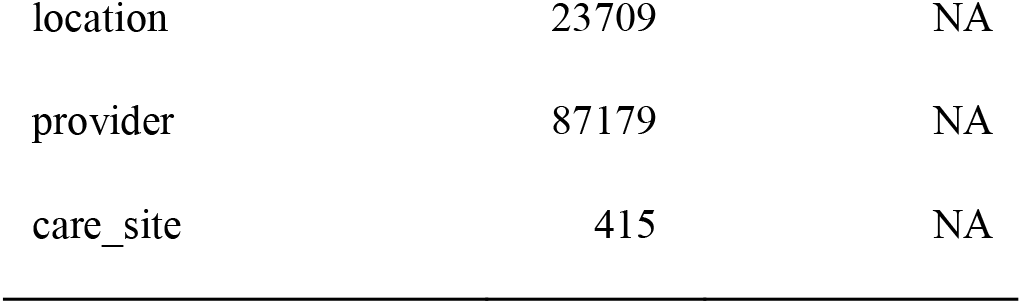
Mapping Summary by Table.

Of the 1164 registry-identified cases, 815 were analytic cases. Among the 815 analytic cases, 26 were incomplete and 89 were discordant. As a result, 700 (85.9%) patients were complete and concordant. Upon manual review of the 26 incomplete cases, we found that 24 patients had a reportable prostate cancer diagnosis (either confirmed or suspicious). However, no corresponding ICD codes were documented in the EHR. The remaining two cases were identified from scanned reports originating from outside institutions. For the 89 patients with discordant diagnosis dates, the results were mixed. Although the diagnosis date in the GCR was accurate for most patients (56; 62.9%), we identified 33 cases (37.1%) in which the GCR diagnosis date was inaccurate. We reported these cases to the cancer registrar to correct the GCR errors. Overall, manual review indicated that most discrepancies in the UTMB-OMOP were attributable to documentation issues, typically leading to a delayed diagnosis date in the UTMB-OMOP relative to the GCR. Despite these, the UTMB-OMOP accurately captured 700 of the 815 analytic patients reported to the GCR. Among these 700 patients, the median age at initial diagnosis was 67.03 years (interquartile range [IQR]: 61.14-72.19 years).

For the PSA values, we identified 673 patients (96.1%) who had at least one PSA measurement in the UTMB-OMOP. The average number of PSA measurements per patient was 12.3 (SD: 10.7). The median PSA follow-up time was 6.8 years (SD: 6.6 years). There were 382 patients diagnosed in or after 2018. This cohort served as the comparison group for PSA, Gleason score, and cancer stage analyses. After restricting the measurement dates to the period from 30 days before diagnosis through the earlier of the initial treatment date recorded in GCR or 365 days after diagnosis (365 days after diagnosis if no treatment was documented), we identified 219 patients (57.3%) with at least one PSA measurement in the UTMB-OMOP, whereas 364 patients (95.3%) had a PSA value recorded in the GCR. Completeness agreement was poor (agreement = 0.615, 95% CI 0.564-0.664). Upon manual review of patients with PSA data in the GCR but not in the UTMB-OMOP, we found that these patients had external (non-UTMB) PSA measurements. These values were not available for data extraction because they were not exported to the SQL database. For patients with PSA measurements in both data sources, the value agreement was excellent (ICC = 0.942, 95% CI 0.925-0.955). After applying the NLP algorithm, the availability agreement became moderate (Agree=0.736, 95% CI 0.688 - 0.779) and value agreement was excellent (ICC = 0.960, 95% CI 0.950-0.969).

We found that the cancer stage table was underutilized in the UTMB-OMOP. There were only 7 Gleason scores extracted from the UTMB-OMOP, whereas the GCR had 382 (100%). We skipped the comparison metrics due to the extremely low sample size. After the NLP extraction, there were 320 (83.8%) Gleason scores. The availability agreement was good (agreement = 0.885, 95% CI 0.848-0.915) and the value agreement was excellent (ICC = 0.969 95% CI 0.962-0.975).

For clinical staging, only 10 patients in the UTMB-OMOP had a cancer stage group versus 331 (86.6%) in the GCR. After NLP extraction, cancer stage groups were identified for 150 (39.3%) patients from UTMB-OMOP notes. The availability agreement was poor (agreement = 0.463, 95% CI 0.412-0.515) but the value agreement was excellent (κ = 0.865 95% CI 0.781-0.950). We manually reviewed all patients without cancer stage groups in the UTMB-OMOP and identified two major reasons. First, some patients had TNM staging information documented at external institutions. Therefore, these data were not transferred into the UTMB-OMOP. Second, some patients did not have TNM staging explicitly documented in the clinical notes. Instead, informal language such as “Has metastatic disease” was used to imply a clinical M1 stage. Our prompt intentionally avoids such language to reduce incorrect inference of staging.

Table 2 summarizes the treatment modality for prostate cancer patients in the UTMB-OMOP and the GCR. In the UTMBOMOP, a total of 90 patients (12.9%) received RP, 119 patients (17%) received RT, and 173 patients (24.7%) received ADT. Compared to the GCR, the completeness agreement was good for RP (agreement=0.896, 95% CI 0.871-0.917), RT (agreement=0.861, 95% CI 0.834-0.886), and ADT(agreement=0.864, 95% CI 0.837-0.889). We applied the missing treatment detection algorithm and identified an additional 36 patients with RP and 28 patients with RT. The completeness agreement stayed good for RP (agreement=0.906, 95% CI 0.882-0.926) and RT (agreement=0.821, 95% CI 0.791-0.849). We manually reviewed the 62 patients flagged by the missing treatment detection algorithm and found that treatment records for 43 patients could be confirmed as accurate. The major source of error was focal therapy (n=9), which is an emerging but not yet standard treatment for prostate cancer patients. Six patients had no treatment records. Four patients were misclassified (either from RP to RT or vice versa).

**Table 2.** Comparison of Treatment Modality Across Data Sources: OMOP: UTMB-OMOP; GCR: Galveston Cancer Registry; Algorithm: UTMB-OMOP combined with with treatment detection algorithm; RP: radical prostatectomy; RT: radiation therapy; ADT: androgen deprivation therapy.

|  | OMOP | GCR | Algorithm |
| --- | --- | --- | --- |
| RP | 90 | 127 | 126 |
| RT | 119 | 126 | 147 |
| ADT | 173 | 166 | 173 |
| Any | 300 | 336 | 364 |

By applying the BCR recurrence algorithm, we identified 60 patients with BCR (22 patients after RP and 38 after RT). The median time to BCR was 3.05 years (IQR: 1.88 - 5.47 years). We manually reviewed all BCR patients. Of the initial 60 patients with BCR of prostate cancer, 50 underwent subsequent clinical evaluation (17 PET, 14 MRI, 16 CT, 2 bone scan, and 1 biopsy). Among these, the recurrence rate was high, with 29 patients having clinically confirmed recurrence or metastasis of prostate cancer (PPV = 48.3%).

## 3 DISCUSSION

We developed and evaluated an OMOP-standardized prostate cancer database at UTMB by transforming UTMB Epic EHR data into the OMOP CDM and assessing data quality across the prostate cancer care continuum. Core OMOP tables were mapped with high completeness.

Comparisons with the GCR showed that most analytic cases had complete and concordant diagnosis information within a four-month window. PSA values, Gleason score, and cancer stage group demonstrated excellent value agreement when present in both sources. Generally speaking, the registry provided more complete data around diagnosis and initial treatment, while the UTMB-OMOP offered richer longitudinal PSA follow-up. The 6-year average follow-up time for the PSA value indicates an excellent longitudinal tracking of prostate cancer patients beyond typical GCR reporting windows.

Several important limitations of the UTMB-OMOP database became clear. First, not all prostate cancer diagnoses receive an ICD code, likely because definitive diagnosis often depends on biopsy results that return after the visit. Moreover, cancer staging and Gleason score are poorly and incompletely captured in structured fields, with most information residing in unstructured notes and pathology reports. In addition, cancer care provided at external facilities is not stored in structured EHR fields (although visible via the Epic interface). Finally, there is no specific code to indicate the first line of treatment or cancer recurrence, which makes it difficult to determine clinical versus pathologic cancer staging and long-term surveillance of treatment outcomes.

To mitigate these issues, we used an LLM to extract stage and Gleason score from unstructured text. To ensure data accuracy, we designed our prompt conservatively and extracted only values explicitly mentioned in the clinical notes. No inference was performed by the LLM (e.g., “Has metastatic disease” was not translated to M1). Therefore, there remains room for further improvement. We also implemented PSA-based algorithms to infer missing RP and RT and to identify BCR. The results showed that these methods greatly improved completeness of our data and value agreement with the cancer registry. However, it should also be noted that the additional algorithms also increase the risk of introducing new data errors (e.g., focal therapy can be flagged by the treatment detection algorithm) and require careful validation. For all comparisons, we specified the time window around diagnosis based on urologists’ input, thereby reducing the risk of using pathologic data in the comparison.

We also identified several data quality issues originating from the GCR (e.g., incorrect diagnosis date). This contradicted the common assumption that cancer registry data are always cleaner. One contributing factor is that the GCR only collects and cleans data from UTMB. It relies on the Texas Cancer Registry to perform the final round of data harmonization. In addition, a 2018 vendor change in the GCR introduced another potential source of error. Our manual review observed that most erroneous cases occurred before 2018, with better data quality in and after 2018. We reported the erroneous data elements to the GCR. Thus, our UTMB-OMOP data can also support improving data quality in the cancer registry.

The study has several limitations. First, the pipeline was evaluated only at a single center (UTMB); variations in Epic EHR configurations across healthcare systems may require site-specific adaptations. Second, successful ETL into the OMOP CDM does not guarantee underlying data quality, which depends on local clinical documentation practices. Third, the cohort was identified through GCR, and corresponding EHR records were extracted using registry-provided MRNs. Thus, the study assessed data completeness and concordance among registry-identified patients, rather than the ability of OMOP data to independently ascertain prostate cancer cases. Patients present in the EHR but absent from GCR were not identified under this design, and the completeness of case ascertainment could not be evaluated. Fourth, although our manual review of sampled reports found no incorrect values among the reviewed positive NLP extractions, some information present in the reports was not extracted. This study did not formally evaluate or validate the LLM prompt used for information extraction. Therefore, the results should be interpreted with caution. Last, manual chart review was conducted by a single primary reviewer with an attending urologist for uncertain cases, rather than independent duplicate assessment. Therefore, interrater agreement was not assessed. The reported kappa and ICC statistics describe agreement between data sources, not between human reviewers. Patients without recurrence were not systematically reviewed due to the large number of patients, so false-negative rates and sensitivity could not be estimated from the chart review.

In the future, we aim to develop an LLM-assisted data extraction pipeline for cancer data extraction. This tool will help ensure data quality and reduce labor costs. In addition, we will use the existing UTMB-OMOP to build a machine learning model for early detection of cancer recurrence. This will facilitate cancer risk assessment and early screening for patients with prostate cancer. With the rich longitudinal data available in UTMB-OMOP, we expect that the resulting machine learning model will be more capable compared to models built with cross-sectional data (e.g., data from cancer registries).

Overall, we successfully built a UTMB-OMOP data pipeline. The incorporation of PSA-based algorithms and NLP substantially improved the completeness and utility of prostate cancer data at UTMB. This work offers a reproducible model for other institutions seeking to standardize EHR data for prostate cancer research.

## Supporting information

Supplemental sections

## Data Availability

The data produced in this study will not be available online. However, the authors welcome external collaboration request.

## 4 CODE AVAILABILITY

Public sharing of the full ETL pipeline is restricted because it contains nonpublic Epic schema elements. The data pipeline code can be found on the Epic UserWeb forum https://userweb.epic.com/Thread/151097/An-R-pipeline-from-Clarity-toOMOP-CDM/. A general description provided in Supplemental section 2. The PSA-based SIGDROP algorithm is available on GitHub https://github.com/Jiefei-Wang/SIGDROP.

## 5 Funding Statement

This work was supported by the Data Management and Analysis Core (DMAC) at the University of Texas Medical Branch (UTMB), which is funded by the Cancer Prevention and Research Institute of Texas (CPRIT; Grant number: RP210130)

## 6 Data Availability Statement

Because the data used in this study contain protected health information, they cannot be shared publicly. Readers interested in using these data can contact the corresponding author to discuss potential collaboration.

## 7 Contributorship Statement

Jiefei Wang developed the data pipeline, conducted the data analysis, and interpreted the study results. Jamaal C. Jackson contributed to the conceptual design of the study, supervised clinical chart review, provided clinical adjudication and data verification for ambiguous cases, and reviewed the manuscript for clinical accuracy. Anette Garza contributed to the development of natural language processing methods and reviewed patient data. Suhas Nalla served as the primary clinical chart reviewer under the supervision of Jamaal C. Jackson. Tammy Adams Ninnemann reviewed patient data and provided expertise in cancer registry practices. Yuanyi Zhang contributed to the development of the data pipeline. Yong-Fang Kuo reviewed the manuscript and validated the analytical methods and study findings.

## 8 Conflict of Interests Statement

The authors declare no conflicts of interest.

## Notes

### Competing Interest Statement

The authors have declared no competing interest.

### Author Declarations

The University of Texas Medical Branch Institutional Review Board <> has approved this study. Protocal Number: 23-0223

### Summary of Updates

revise based on reviewer comments.

