## Supplemental sections for "Developing an OMOP-Standardized Prostate Cancer Database and Improving Data Quality Using NLP and PSA-Based Algorithms"

### 1 OMOP Table Definitions

Table 1: Core OMOP CDM v5.4 Tables

| **Name** | **Description** |
| --- | --- |
| person | A unique patient with demographic information such as sex, date of birth, and race. One row per patient. |
| condition_occurrence | Diagnoses, signs, or symptoms recorded for a patient. Each row represents a single condition record for a patient. |
| measurement | Quantitative or qualitative clinical measurements (e.g., laboratory values, vital signs). Each row represents a single measurement record for a patient. |
| drug_exposure | Exposure to medications, vaccines, or other drug products. Each row represents a single drug exposure record for a patient. |
| procedure_occurrence | Procedures or interventions performed for diagnostic or therapeutic purposes. Each row represents a single procedure record for a patient. |
| visit_occurrence | A healthcare encounter (e.g., inpatient, outpatient, emergency visit). Each row represents a single visit for a patient. |
| observation | Clinical or contextual facts not captured in other domains (e.g., alcohol use, family history). Each row represents a single observation record for a patient. |
| death | Information about a patient’s death, including date and cause (if available). One row per patient. |
| note | Unstructured clinical text recorded for a patient. Each row represents a single clinical note for a patient. |
| location | Physical address information for persons or care sites. |
| provider | A healthcare professional involved in care delivery. |
| care_site | A healthcare facility where care is delivered. |

### 2 Epic Clarity to OMOP ETL Mapping

Table 2: Representative Epic source tables, transformation rules, and destination OMOP CDM tables. The source table list is not exhaustive and includes only tables publicly documented or referenced in Epic’s EHI documentation (<https://open.epic.com/EHITables/TechSpec>, © 2026 Epic Systems Corporation.). A single Epic table may populate multiple OMOP tables, depending on the domain of the mapped standard concept.

| **Epic Domain** | **Epic source tables** | **Transformation rules** | **OMOP tables** |
| --- | --- | --- | --- |
| Demographics | PATIENT, CLARITY_POS | Extract demographics, locations, and deaths; Assign person_id to each patient; All other OMOP tables should be linked to the person table via the person_id column. | person, location, death |
| Providers and care locations | CLARITY_SER, CLARITY_LOC,  CLARITY_DEP | Assign provider_id to each provider and link providers, locations, and care sites. | provider, location, care_site |
| Encounters | PAT_ENC, PAT_ENC_HSP | Prefer hospital records for shared encounter IDs. | visit_occurrence |
| Diagnoses | PAT_ENC_DX | Map ICD-9/10 codes to standard concepts, preferring ICD-10; route by concept domain. | condition_occurrence, measurement, observation, procedure_occurrence |
| Laboratory results | ORDER_PROC, ORDER_RESULTS | Map lab codes; parse values and operators; retain source units. | measurement, observation |
| Medication orders and administrations | ORDER_MED, ORDER_DISP_MEDS | Map RxNorm/NDC codes; distinguish prescriptions from administrations. | drug_exposure |
| Procedure orders and surgery | ORDER_PROC, OR_LOG_ALL_PROC | Map CPT4, ICD-9-CM procedure and ICD-10-PCS codes to standard concepts; align dates and encounter links. | procedure_occurrence |
| Clinical notes | HNO_INFO, HNO_NOTE_TEXT, RES_ DB_MAIN, SPEC_DB_MAIN | Combine note text; exclude missing/empty text. | note |
| Social history | SOCIAL_HX | Convert history fields to observations (e.g., smoking, alcohol use). | observation |

We developed an Extract, Transform, and Load pipeline to convert Epic EHR data from the University of Texas Medical Branch (UTMB) into the OMOP CDM v5.4 format. Implemented in R (v4.2), the pipeline utilized tidyverse for data transformation, dbplyr for remote database queries, and targets for workflow orchestration. Table 2 summarizes the mappings between Epic Clarity and OMOP CDM tables.

We identified the study cohort using medical record numbers (MRNs) from the UTMB Galveston Cancer Registry and extracted their corresponding Clarity records, including patient demographics, encounters, diagnoses, procedures, medications, and laboratory and clinical measurements. Source queries were filtered on patient MRN or internal identifier to restrict data volume to the target cohort.

To interpret source data, we extracted Epic reference tables (ZC tables), which provide descriptions for local categorical values and mappings to clinical coding systems such as ICD-9-CM and ICD-10-CM. OMOP vocabularies and concept relationships were retrieved from Athena to map source terms to standard OMOP concepts.

Each dataset was transformed to conform to the OMOP schema by standardizing field names, aligning data types, and routing records to destination tables by clinical domain (e.g., condition, procedure, drug, or measurement). Consequently, individual source tables could populate multiple OMOP tables. Transformed records were then consolidated into their target tables, assigned synthetic primary keys, and linked via foreign keys.

Finally, We mapped institution-specific attributes (e.g., patient gender and provider specialties) to standard OMOP concepts. Any remaining unmapped records were assigned concept ID 0. The resulting tables were stored in DuckDB for downstream analysis. We generated two database versions: a strictly compliant OMOP CDM instance and an extended version retaining source identifiers (e.g., patient MRN) for complete end-to-end traceability.

### 3 NLP Data Extraction Pipeline

We used the Gemma 4 26B model from Hugging Face (RedHatAI/gemma-4-26B-A4B-it-FP8Dynamic) to identify prostate cancer staging information from clinical notes. Notes were first linked to the provider table and filtered to specialties most likely to document staging information. We included specialties related to urology, oncology, pathology, and radiology. For each note, the note date, title, and full text were combined and submitted to the model using a standardized prompt (see Section 4) to extract predefined variables according to a structured output schema. The output schema was defined using Pydantic and inserted into the prompt template at runtime. The prompt also instructed the model to return the exact supporting text span from the note, the closest relevant date, and a confidence level for each extracted finding.

During the prompt development, Anette Garza, a master’s student, reviewed all low- and medium-confidence NLP outputs. We also randomly sampled 100 reports with at least one extracted values for each category (PSA, Gleason score, and TNM stage) for manual review. We iteratively refined prompt until the extracted value were correct. Note that we only evaluated the accuracy of the extracted values, not the completeness of the extraction. The goal of the prompt development was to ensure that the model could accurately extract values without hallucinating. It is possible that the model may have missed some values.

After extraction, we cleaned and normalized each variable set before analysis. For time-sensitive variables such as Gleason scores and PSA values, we kept only records with complete day-level dates and excluded records with dates occurring after the note date. Gleason values were additionally checked for internal consistency and valid ranges: primary and secondary patterns were restricted to 3 to 5, total score to 6 to 10, and grade group to 1 to 5; when the total score or grade group was missing, these were derived from the primary and secondary patterns, and discordant or implausible records were removed (e.g. 3+3=7; total Gleason score = 9 but grade group = 3). PSA values were converted to numeric form and restricted to the standard unit of ng/mL (all PSAs with nonstandard units were excluded). TNM extractions were normalized by parsing T, N, and M strings into standardized components, including stage prefix, numeric stage, and subtype.

We followed AJCC 8th edition rules in Table 3 to define stage group. For each patient, candidate staging variables were restricted to a diagnostic window spanning 30 days before through 120 days after the prostate cancer diagnosis date. Within that window, we found the clinical T, N, and M components that were closest to diagnosis for each axis; unknown extracted stage values coded as X were treated as 0 for N and M when appropriate. Pretreatment Gleason grade group was defined as the maximum grade group observed in the same diagnostic window before treatment, and pretreatment PSA was defined as the maximum PSA value in that window. These components were then mapped to numeric clinical stage groups.

### 4 Prompt Design

You are a clinical data extraction assistant. Your task is to read a medical note and extract specific information related to prostate cancer staging.

Find all distinct mentions of:

- Gleason score
- TNM
- Stage group
- PSA

Use this output schema exactly:

{schema}

Rules:

- Return JSON only.
- One object per distinct mention.
- Keep the clearest instance if the same mention is repeated.
- If nothing is found, return empty lists for all fields.
- If the report is not for prostate cancer, return empty lists for all fields.
- Do not infer missing values, even if they are easy to calculate.
- Use null for any field that is not explicitly stated.
- Use JSON null, not Python None.
- For dates, capture the closest relevant date:
- If the finding has its own nearby date, use that date.
- If no finding-specific date is present and the finding is clearly from the current note, use the note date.
- If the finding is clearly historical and no date is given, use null.
- For date_text, copy the date exactly as it appears in the note, or use null when no date is stated.
- If only a year is given, set date_precision to "year" and date_month and date_day to null.
- If year and month are given but no day, set date_precision to "month" and date_day to null.
- If year, month, and day are all given, set date_precision to "day".
- Year is always the 4-digit year if available
- For supporting_text, always copy a literal supporting span from the note. Do not correct typos or standardize wording. Do not include date as it is captured separately.
- Use confidence:
- high: explicit, clear mention
- medium: likely staging finding but context is somewhat ambiguous
- low: weak or uncertain evidence

Gleason score rules:

- Extract primary, secondary, tertiary, total, and grade_group only when explicitly stated.
- For "Gleason score 7 (4+3)", extract total=7, primary=4, secondary=3.
- For "Gleason 4+3", extract primary=4 and secondary=3, but do not infer total unless explicitly stated.
- For "Grade Group 3", extract grade_group=3.
- For "Gleason score of 6", extract total=6 but do not infer primary or secondary.

TNM rules:

- Extract T, N, and M components when stated together or separately.
- Preserve clinical/pathologic prefixes when present, such as cT2a or pT3b.
- Use stage_basis="clinical" only when explicitly clinical or prefixed with c.
- Use stage_basis="pathological" only when explicitly pathologic/pathological or prefixed with p.
- If TNM components have mixed prefixes, preserve each component as written and set stage_basis only if the overall basis is explicit.

Stage group rules:

- Extract prostate cancer stage groups such as Stage 2, Stage II, Stage IIB, AJCC stage IV, and clinical stage IIIA. Ignore TNM staging; it should be captured in the TNM section.
- Put only the number in main_group.
- Put only the suffix letter in subgroup.
- Set subgroup=null if there is no suffix.
- Set is_ajcc=null if the staging system is not stated.
- Set ajcc_edition=null if the edition is not stated. If AJCC is stated but no edition, set ajcc_edition=null.

PSA rules:

- Extract only PSA values that are prostate-specific antigen results.
- Capture the numeric value and normalize unit to one of: "ng/mL", "ug/dL", or "pg/mL".
- If the unit is not stated, use null. If the unit is stated but vague (e.g. NG), use null.
- Ignore unrelated abbreviations or non-lab uses of "PSA".
- Ignore "elevated PSA".
- Ignore free PSA percentage.

Now extract from this note:

```txt

{note_text}

```

### 5 AJCC Cancer Stage Group

Table 3: Prostate Cancer Staging AJCC

| **Stage** | **T (Tumor)** | **N (Node)** | **M (Met)** | **PSA Level** | **Grade Group** |
| --- | --- | --- | --- | --- | --- |
| Stage I | cT1, cT2a, or pT2 | N0 | M0 | *<* 10 | 1 |
| Stage IIA | cT1, cT2a, or pT2 | N0 | M0 | 10 to *<* 20 | 1 |
|  | cT2b or cT2c | N0 | M0 | *<* 20 | 1 |
| Stage IIB | T1 or T2 | N0 | M0 | *<* 20 | 2 |
| Stage IIC | T1 or T2 | N0 | M0 | *<* 20 | 3 or 4 |
| Stage IIIA Stage IIIB | T1 or T2  T3 or T4 | N0  N0 | M0  M0 | ≥ 20  Any PSA | 1 to 4  1 to 4 |
| Stage IIIC | Any T | N0 | M0 | Any PSA | 5 |
| Stage IVA | Any T | N1 (Regional) | M0 | Any PSA | Any Grade |
| Stage IVB | Any T | Any N | M1 (Distant) | Any PSA | Any Grade |

### 6 PSA-based Detection of Potentially Missing Treatments

We implemented the SIGDROP/DTX pseudocode of Batouche et al.[1] in R. Algorithms 1 and 2 specify the implementation used in this study. We used the same significant-decline thresholds and the RP/RT cutoff. Missing PSA was excluded from the analysis. Three modifications were made to the original pseudocode: (1) we saved each first threshold-crossing decline and restarted the peak search, allowing multiple candidate declines per patient rather than returning a single result after scanning the PSA series; (2) we explicitly excluded candidates associated with recorded RP, RT, or ADT, extending the exclusion interval by 30 days before the peak and after the qualifying measurement; and (3) we bounded the denominator of the relative PSA decline calculation below by 0.0001 to prevent division by zero.

For treatment comparisons, inferred and recorded treatments were combined and restricted to 30 days before through 365 days after diagnosis. The earliest record per patient and treatment type was retained; patients were counted once per modality and once overall for any treatment.

**Algorithm 1** Study implementation of SIGDROP: candidate PSA declines

**Require:** Date-ordered PSA values $p_{1}, \ldots, p_{n}$ and dates $d_{1}, \ldots, d_{n}$

**Ensure:** List $C$ of tuples $(D, N, P, m)$; empty if no candidate

1: $C \leftarrow[]$
2: **if** $n < 2$ **then**
3: **return** $C$
4: **end if**
5: $P \leftarrow p_{1}; D \leftarrow d_{1}$
6: **for** $j = 1, \ldots, n - 1$ **do**
7: $\delta\leftarrow d_{j+1} - D; e \leftarrow p_{j} - p_{j+1}$
8: **if** $e \leq0$ **then**
9: **if** $p_{j+1} \geq P$ **or** $\delta> 365$ **then**
10: $P \leftarrow p_{j+1}; D \leftarrow d_{j+1}$
11: **end if**
12: **else**
13: $\beta\leftarrow P - p_{j+1}; \alpha\leftarrow\beta/max(P, 0.0001)$
14: **if** $(\alpha\geq0.75 and \beta\geq3)$ **or** $(\alpha\geq0.50 and \beta\geq4)$ **then**
15: Append $(D, d_{j+1}, P, p_{j+1})$ to $C$
16: **if** $j + 2 > n$ **then**
17: **break**
18: **end if**
19: $P \leftarrow p_{j+2}; D \leftarrow d_{j+2}$
20: **else**
21: **if** $\delta> 365$ **then**
22: $P \leftarrow p_{j+1}; D \leftarrow d_{j+1}$
23: **else if** $j + 2 \leq n$ **then**
24: **if** $d_{j+2} - D > 365$ **then**
25: $P \leftarrow p_{j+1}; D \leftarrow d_{j+1}$
26: **end if**
27: **end if**
28: **end if**
29: **end if**
30: **end for**
31: **return** $C$

**Algorithm 2** Study implementation of DTX: potentially missing treatments

**Require:** Patients’ PSA series and recorded RP, RT, and ADT dates

**Ensure:** List $L$ of inferred treatment records

1: $L \leftarrow[]$
2: **for** each patient $i$ **do**
3: $C_{i} \leftarrow$SIGDROP$(p_{i}, d_{i})$ using Algorithm 1
4: **for** each $(D, N, P, m)$ in $C_{i}$ **do**
5: **if** no recorded RP, RT, or ADT date $t$ satisfies $D - 30 < t < N + 30$ **then**
6: **if** $m < 0.1$ **then**
7: $q \leftarrow$RP
8: **else**
9: $q \leftarrow$RT
10: **end if**
11: Append $(i, q, D)$ to $L$
12: **end if**
13: **end for**
14: **end for**
15: **return** $L$

### References

[1] Abderrahim Oussama Batouche, Eugen Czeizler, Miika Koskinen, Tuomas Mirtti, and Antti Sakari Rannikko. Synergizing data imputation and electronic health records for advancing prostate cancer research: Challenges, and practical applications. *arXiv preprint arXiv:2311.02086*, 2023.
